# Body Fatness Increases Risk of Monoclonal Gammopathy of Undetermined Significance in the U.S. Population

**DOI:** 10.1101/2024.10.10.24314907

**Authors:** Mengmeng Ji, John Huber, Mei Wang, Yi-Hsuan Shih, Tuo Lan, Shi-Yi Wang, Su-Hsin Chang

## Abstract

**Objective:** Current evidence on whether obesity is associated with MGUS remains inconsistent. This study aims to evaluate the association between objectively measured obesity markers and the risk of developing MGUS, using nationally representative data from the U.S. population.

**Method:** Data came from the third National Health and Nutrition Examination Survey III (1988-1994) and continuous NHANES (1999-2004). Multivariable-adjusted odds ratios (aOR) and 95% confidence intervals (CIs) for the association between the risk of developing MGUS and seven obesity markers—including baseline body mass index (BMI), maximum lifetime BMI, waist circumference (WC), waist-hip ratio (WHR), total body fat, fat-free mass, and body percentage fat—were estimated using logistic regression.

**Results:** In the study cohort, a total of 364 participants tested positive for MGUS, compared to 12,043 participants without MGUS. Multivariate logistic regression analysis indicates that each 1% increase in body fat percentage is associated with a 4% (aOR: 1.04, 95% CI [1.01, 1.07]) higher risk of MGUS. Each 1% increase in body fat percentage is also associated with a 6% (aOR: 1.06, 95% CI [1.02, 1.10]) higher risk of non-IgM MGUS. No statistically significant association was found between MGUS and other obesity markers, including baseline BMI, maximum lifetime BMI, WC, WHR, and fat-free mass.

**Conclusion:** Our findings show that obesity is associated with an increase in the risk of MGUS. However, many obesity markers, including the commonly used BMI, fail to capture this association.

## INTRODUCTION

Monoclonal gammopathy of undetermined significance (MGUS) is an asymptomatic plasma cell disorder that overproduces serum monoclonal protein and could progress to multiple myeloma (MM), a fatal cancer of plasma cells.^1,2^ MGUS is one of the most prevalent premalignant conditions^3,4^; however, due to its asymptomatic nature, research on risks associated with MGUS is relatively limited despite its high prevalence.^5^ The present evidence suggests that older age, male sex, black race, prior infections and inflammatory conditions, and a family history of a plasma cell malignancy increase the risk of developing MGUS.^4-6^

Obesity is concluded to be the only modifiable factors for MM risk^7^ and also plays a role in the progression of MGUS to MM.^8,9^ But findings on whether obesity is associated with MGUS remain inconsistent.^5,10,11^ Population-based studies of Reykjavik and of Heinz Nixdorf reported null association between obesity (as measured by 11 obesity markers) and MGUS.^12,13^ A study using a large primary-care database of UK also reported null association between BMI and MGUS.^14^ However, two studies conducted in the U.S. reported positive association between obesity and MGUS.^15,16^ One study was based on Southern Community Cohort Study (SCCS) cohort and found that self-reported body mass index (BMI) were independently associated with an 80% higher excess risk of MGUS among women.^15^ A recent study of 2,628 participants in the PROMISE trial, who are at high risk of MM, reported that BMI is positively associated with mass spectrometry-detected MGUS.^16^ The discrepancy between the European and U.S. studies may be attributed to differences in participant selection, the use of subjective measurements, and the use of varying obesity markers.

This study aims to evaluate the association between objectively measured obesity markers and the risk of developing MGUS, using nationally representative data of the U.S. population. The findings of this study could advance the understanding of risk factors associated with MGUS and provide implications for potential MGUS screening programs.

## METHOD

### Data and Study Population

Data came from the third National Health and Nutrition Examination Survey III (1988-1994) and continuous NHANES 1999-2000, 2001-2002, 2003-2004. NHANES used a stratified multistage probability cluster design to draw samples representative of the civilian, non-institutionalized U.S. population.^17,18^ The NHANES survey combines household interviews and physical examinations, including physical and physiological measurements, blood collection and laboratory tests.

The NHANES was approved by the National Center for Health Statistics (NCHS) Research Ethics Review Board, and all participants provided written informed consent. This study utilized de-identified NHANES data that is publicly available and was therefore deemed exempt from human subjects review.

### MGUS Diagnosis

Testing for the presence of MGUS in NHANES was performed at the Protein Immunology Laboratory at Mayo Clinic, Rochester, Minnesota.^19^ First, serum samples were analyzed for all identified subjects by conventional agarose-gel electrophoresis to reveal the occurrence and pattern of monoclonal protein in the study cohort. Samples with an equivocal or definite M-protein present on electrophoresis were then subjected to serum protein immunofixation, and to serum-free light-chain assay for confirmation and typing of the M-protein. In NHANES III and NHANES 1999-2004, 6,560 and 5,847 participants≥ 50 years of age were screened for MGUS using serum samples, respectively.

### Obesity Markers

Participants were weighed in mobile examination centers, wearing only underclothing and an examination gown. Weight was recorded on a digital scale in kilograms. Standing height was measured using a stadiometer with a fixed vertical backboard and an adjustable headpiece. Waist circumference (WC) was measured just above the iliac crest using a steel measuring tape. The NHANES anthropometry procedures manual provides detailed descriptions of the measurement protocol, equipment, and quality control.^20^ BMI was calculated as measured weight in kilograms divided by squared height in meters. The classifications were underweight (<18.5 kg/m^2^), normal weight (18.5-24.9 kg/m^2^), overweight (25.0-29.9 kg/m^2^), and obese (>30.0 kg/m^2^). Abdominal obesity was defined as a WC of at least 102 cm for men and at least 88 cm for women. Waist–hip ratio (WHR) was calculated dividing waist measurement by hip measurement. In addition, maximum lifetime BMI was constructed by using self-reported maximum lifetime weight (based on an NHANES question that asks participants to recall maximum weight) and subjectively measured height.

Body composition, including total body fat (TBF) (kg), fat-free mass (FFM) (kg), body percentage fat (%BF) was measured for adults aged 8 years and over (pregnant females excluded) by using dual energy X ray absorptiometry (DXA) in NHANES 1999-2004.^21^ About 22% of subjects in the NHANES 1999–2004 had at least one missing regional body fat measurement that was due to invalid DXA scanning.^22^ Because DXA data missingness is related to age, BMI, weight and height, and possibly other characteristics, participants with missing data cannot be treated as a random subset of the original sample.^23^ For this reason, the NHANES 1999–2004 generated 5 imputed data sets for missing DXA regional body-composition measurements.^22,23^

In the NHANES III, participants aged 12 years and over (pregnant females excluded) had a single, tetrapolar bioelectrical impedance analysis (BIA) measurement of resistance and reactance at 50 kHz taken between the right wrist and ankle while in a supine position, using Valhalla 1990B Bio-Resistance Body Composition Analyzer (Valhalla Scientific, San Diego, CA, USA).^24^ Following previous studies^25-27^, the established equations listed below were applied to the respective NHANES III anthropometric and converted BIA resistance data to derive estimates for FFM.

Males: FFM = −10.678 + 0.262*Weight + 0.652*(S^2^/Resistance) + 0.015*Resistance (r^2^ = 0.90, RMSE = 3.9 kg),

Females: FFM = −9.529 + 0.168*Weight + 0.696*(S^2^/Resistance) + 0.016*Resistance (r^2^ = 0.83, RMSE = 2.9 kg), where S refers to stature in cm.

Estimates for TBF and %BF for each NHANES III participant were derived from their corresponding estimated FFM using the equations: TBF = weight−FFM; %BF = TBF/weight.

### Covariates

Covariates in the analysis included age at MGUS screening, gender (male/female), race/ethnicity (non-Hispanic Whites, non-Hispanic Blacks, Mexican American, and other race), educational attainment (less than 9th grade education, 9-11th grade education, high school, some college or associates degree, college or higher), and income-to-poverty ratio (low: <1.85, middle: 1.85-3.5, high: >3.5).

### Statistical Analysis

Descriptive statistics including means and standard deviations for estimates of social-demographics and obesity markers were calculated for participants with and without MGUS. All analyses were accounted for pseudo-strata, pseudo-sampling units and participant weights to accommodate the complex sampling of the NHANES, following the analytic guidelines.^28^

Multivariable-adjusted odds ratios (aOR) and 95% confidence intervals (CIs) for the association between the risk of developing MGUS and 7 obesity markers (i.e., baseline BMI, maximum lifetime BMI, WC, WHR, FFM, TBF, %BF) were estimated using logistic regression. Considering the high correlation between different obesity markers (**Supplementary Figure S1**), separate model was conducted for each obesity marker.

The statistical approach for 5 imputed datasets of body composition in NHANES 1999-2004 was to analyze each dataset separately and then combining the estimates and standard errors based on Rubin’s rules for repeated-imputation inference. In brief, the combined estimate of regression coefficient (*Q*) is simply the mean of 5 individual estimates 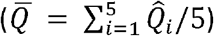. The combined standard error for 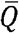 is based on the following quantities: the within-imputation variance 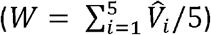 and between-imputation variance 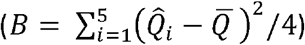, where 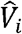 denote the associated variance estimates of the 5 imputed datasets. The total variance combines the within- and between-imputation variances 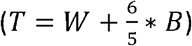 The square root of this total variance, 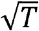, is the combined standard error of the combined estimate 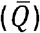. The details were described in the technical documentation by NCHS.^29^

In the sensitivity analysis, only patients with immunoglobulin (Ig) G, IgA or light chain MGUS were included since IgM MGUS typically progresses to lymphoma and rarely to MM and IgD/IgE MGUS is very rare.^30,31^

All tests were two-sided, and p-values <0.05 were considered to be statistically significant. All analyses were conducted with in STATA 17 SE version (StataCorp, College Station, TX).

## RESULTS

In the study cohort, a total of 364 participants were tested with MGUS, compared to 12,043 participants without MGUS. In the MGUS group, the distribution of immunoglobulin (Ig) isotypes was as follows: 9.66% of participants had IgA MGUS, 69.66% had IgG MGUS, 12.80% had IgM MGUS, and 7.88% had biclonal MGUS. Age was higher among those with MGUS (69.23 vs 64.07 years; P < 0.001), and a larger proportion of MGUS cases were male (54.55% vs 45.26%; P = 0.011). Racial differences were observed, with a higher percentage of non-Hispanic Black participants in the MGUS group (13.06% vs 8.06%; P = 0.028). No significant differences were found in educational attainment or poverty income ratio between groups. The mean BMI at screening was similar between the two groups (28.14 vs 27.96; P = 0.606), and there were no significant differences in BMI categories. Maximum lifetime BMI and waist circumference were also comparable between groups (P = 0.363 and P = 0.210, respectively). However, the waist-to-hip ratio was significantly higher in the MGUS group (0.97 vs 0.95; P = 0.032). The summary was presented in **Table 1**.

**Table 1:** Characteristics of the study participants from NHANES III and NHANES 1999-2004.

|  |  |  |  |
| --- | --- | --- | --- |
|  | With MGUS<br>n = 364 | Without MGUS<br>n = 12,043 | P-value |
| Obesity Markers |  |  |  |
| BMI at screening, mean (SE) | 28.14 (0.34) | 27.96 (0.09) | 0.606 |
| BMI categories, % (SE) |  |  |  |
| Normal: 18.5 to 25 | 25.28 (2.74) | 30.11 (0.71) | 0.562 |
| Underweight: ≤18.5 | 1.72 (0.83) | 1.36 (0.15) |  |
| Overweight: 25.0 to <30 | 40.16 (2.90) | 37.30 (0.55) |  |
| Obese: ≥30 | 30.79 (3.00) | 29.40 (0.63) |  |
| Maximum lifetime BMI, mean (SE) | 31.19 (0.59) | 30.66 (0.15) | 0.363 |
| Waist circumference (cm), mean (SE) | 99.73 (0.99) | 98.46 (0.22) | 0.210 |
| Waist categories, % (SE) |  |  |  |
| Normal risk (men ≤102 cm; women ≤88 cm) | 42.04 (3.63) | 40.92 (0.80) | 0.756 |
| High risk (men >102 cm; women >88 cm) | 57.96 (3.63) | 59.08 (0.80) |  |
| Waist to hip ratio<sup>b</sup> | 0.97 (0.01) | 0.95 (0.002) | 0.032 |
| Fat-free mass (kg) | 51.91 (0.90) | 50.94 (0.17) | 0.321 |
| Total body fat (kg) | 27.35 (0.71) | 27.54 (0.19) | 0.788 |
| Body percentage fat (%) | 33.92 (0.52) | 34.65 (0.16) | 0.190 |
| MGUS Ig isotype, n (%) |  |  |  |
| IgA | 26 (9.66%) | / | / |
| IgG | 259 (69.66%) | / |  |
| IgM | 44 (12.80%) | / |  |
| biclonal | 35 (7.88%) | / |  |
| Age in years, mean (SE) | 69.23 (0.68) | 64.07 (0.21) | <0.001 |
| Gender, % (95%CI) |  |  |  |
| Male | 54.55 (3.3) | 45.26 (0.49) | 0.011 |
| Female | 45.45 (3.3) | 54.74 (0.49) |  |
| Race/ethnicity, % (SE) |  |  |  |
| Non-Hispanic White | 79.67 (2.86) | 81.43 (1.29) | 0.028 |
| Non-Hispanic Black | 13.06 (1.75) | 8.06 (0.57) |  |
| Hispanic | 2.4 (0.6) | 3.2 (0.43) |  |
| Other | 4.88 (1.79) | 7.31 (0.96) |  |
| Education, % (SE) |  |  |  |
| <9<sup>th</sup> grade | 15.73 (2.65) | 15.25 (0.64) | 0.946 |
| 9-11<sup>th</sup> grade | 16.16 (2.72) | 14.43 (0.62) |  |
| High school | 28.09 (3.12) | 29.24 (0.68) |  |
| Some college | 20.61 (2.54) | 20.86 (0.62) |  |
| College graduate or higher | 19.41 (2.22) | 20.21 (0.82) |  |
| Poverty income ratio, % (SE) |  |  |  |
| Low (<1.85) | 26.48 (2.71) | 27.29 (1.2) | 0.195 |
| Middle (1.85-3.5) | 33.01 (3.32) | 26.87 (0.74) |  |
| High (>3.5) | 33.68 (3.16) | 37.53 (1.27) |  |
| Missing | 6.84 (1.46) | 8.31 (0.51) |  |

<sup>a</sup>Data came from NHANES 1999-2004 only.
<sup>b</sup>Data came from NHANES III only.
All analyses were accounted for the complex sampling of the NHANES.

Multivariate logistic regression analysis indicates that each 1% increase in body fat percentage is associated with a 4% (aOR: 1.04, 95% CI [1.01. 1.07]) higher risk of MGUS (**Table 2**). The association between fat mass and body fat percentage was more pronounced in the IgG/IgA MGUS subgroup. Each 1% increase in body fat percentage is associated with a 6% (aOR: 1.06, 95% CI [1.02. 1.10]) higher risk of non-IgM MGUS (**Table 3**). No statistically significant association was found between MGUS and other obesity markers including baseline BMI, maximum lifetime BMI, WC, WHR, and fat free mass. The association between body composition and risk of developing MGUS using 5 imputed data from NHANES was presented in **Supplementary Table S1**.

**Table 2:**
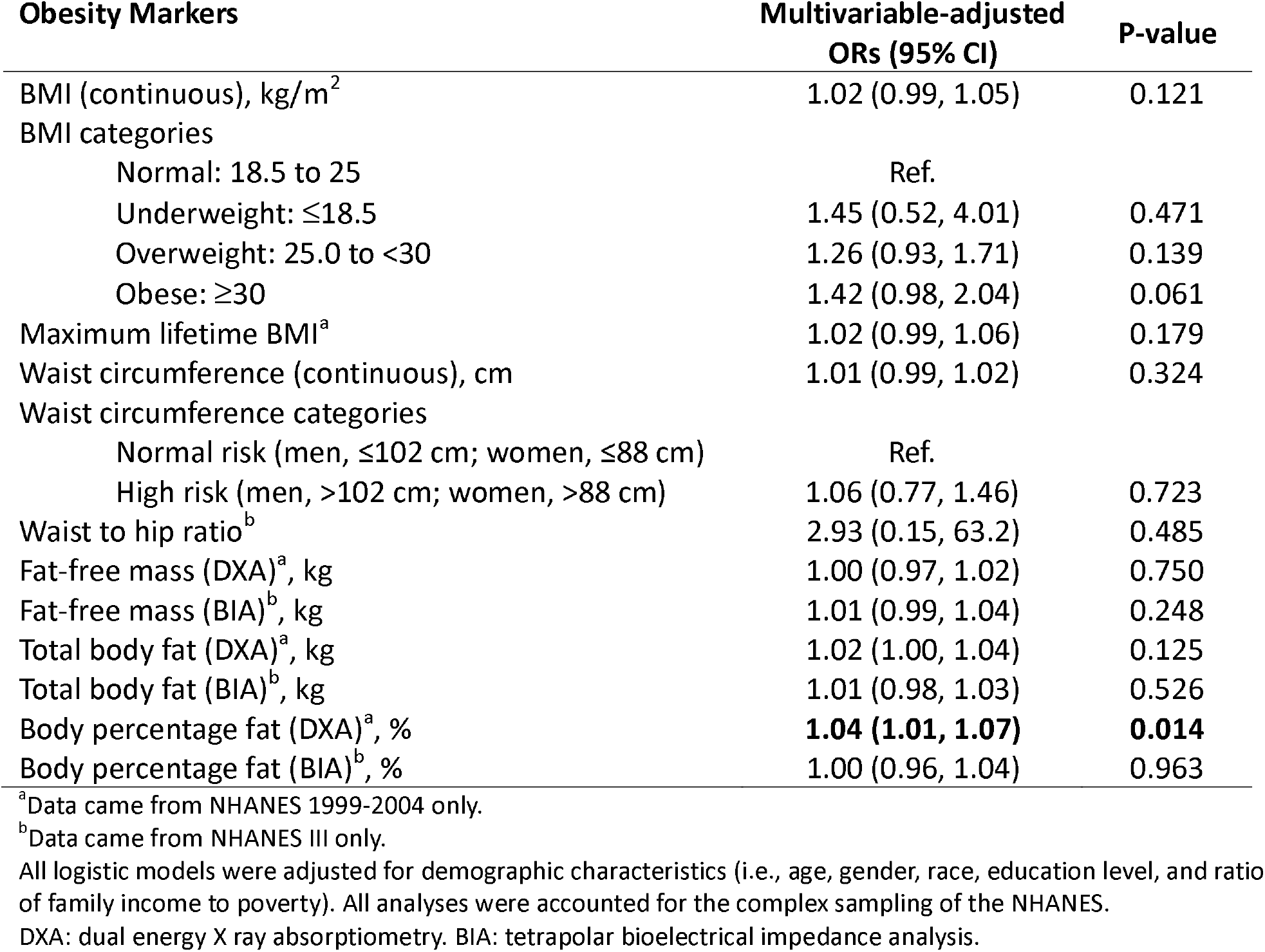
Association between obesity markers and the risk of developing MGUS.

**Table 3:**
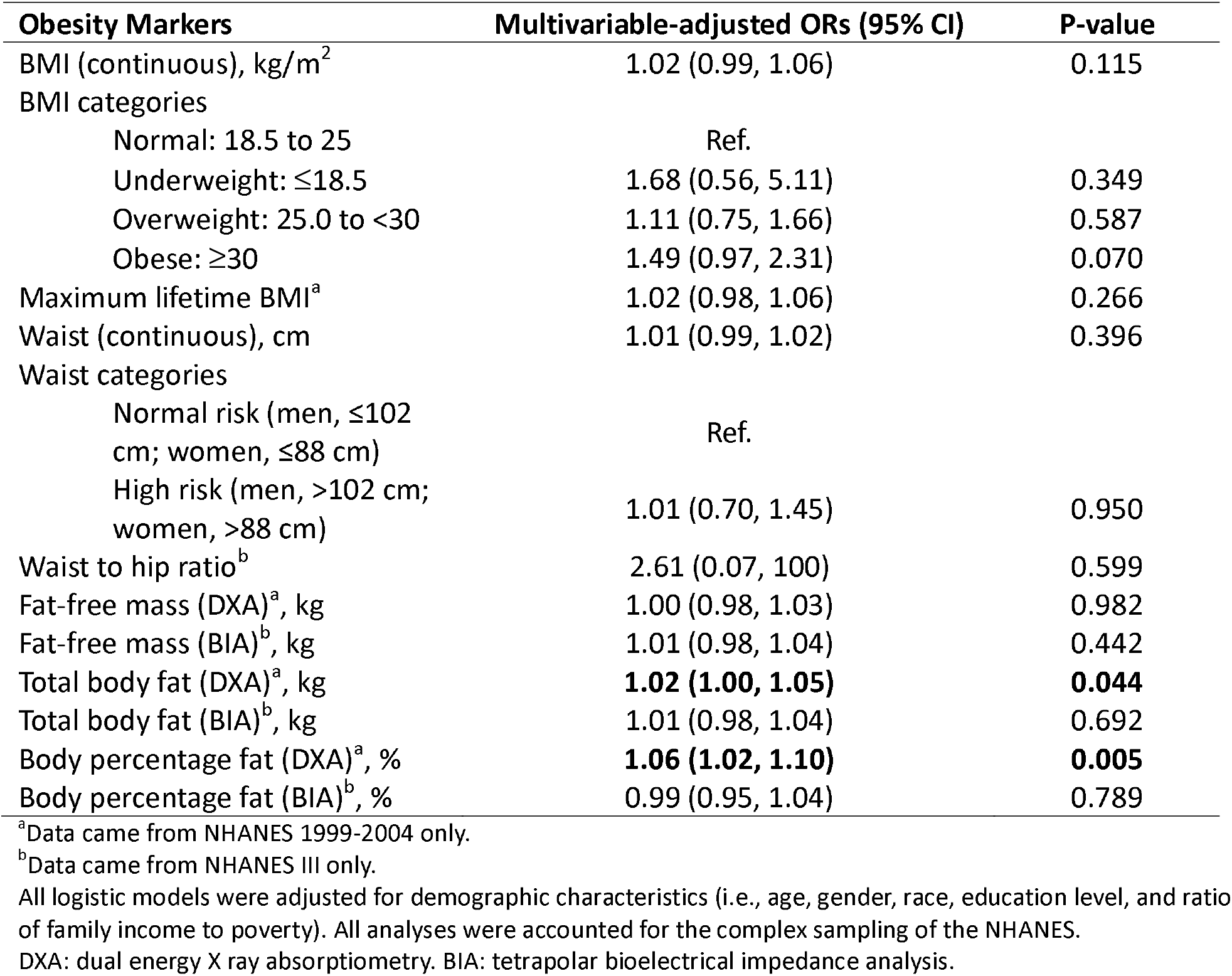
Sensitivity analysis: association between obesity markers and the risk of developing IgG/IgA MGUS.

| **Obesity Markers** | **Multivariable-adjusted ORs (95% CI)** | **P-value** |
| --- | --- | --- |
| BMI (continuous), kg/m<sup>2</sup> | 1.02 (0.99, 1.06) | 0.115 |
| BMI categories |  |  |
| Normal: 18.5 to 25 | Ref. |  |
| Underweight: ≤18.5 | 1.68 (0.56, 5.11) | 0.349 |
| Overweight: 25.0 to <30 | 1.11 (0.75, 1.66) | 0.587 |
| Obese: ≥30 | 1.49 (0.97, 2.31) | 0.070 |
| Maximum lifetime BMI<sup>a</sup> | 1.02 (0.98, 1.06) | 0.266 |
| Waist (continuous), cm | 1.01 (0.99, 1.02) | 0.396 |
| Waist categories |  |  |
| Normal risk (men, ≤102 cm; women, ≤88 cm) | Ref. |  |
| High risk (men, >102 cm; women, >88 cm) | 1.01 (0.70, 1.45) | 0.950 |
| Waist to hip ratio<sup>b</sup> | 2.61 (0.07, 100) | 0.599 |
| Fat-free mass (DXA)<sup>a</sup>, kg | 1.00 (0.98, 1.03) | 0.982 |
| Fat-free mass (BIA)<sup>b</sup>, kg | 1.01 (0.98, 1.04) | 0.442 |
| Total body fat (DXA)<sup>a</sup>, kg | **1.02 (1.00, 1.05)** | **0.044** |
| Total body fat (BIA)<sup>b</sup>, kg | 1.01 (0.98, 1.04) | 0.692 |
| Body percentage fat (DXA)<sup>a</sup>, % | **1.06 (1.02, 1.10)** | **0.005** |
| Body percentage fat (BIA)<sup>b</sup>, % | 0.99 (0.95, 1.04) | 0.789 |

<sup>a</sup>Data came from NHANES 1999-2004 only.
<sup>b</sup>Data came from NHANES III only.
All logistic models were adjusted for demographic characteristics (i.e., age, gender, race, education level, and ratio of family income to poverty). All analyses were accounted for the complex sampling of the NHANES.
DXA: dual energy X ray absorptiometry. BIA: tetrapolar bioelectrical impedance analysis.

## DISCUSSION

This study is the first to utilize nationally representative data to investigate the association between obesity markers and the risk of MGUS in U.S. population. Our findings indicate that a 1% increase in body fat percentage is associated with a 6% increase in the risk of non-IgM MGUS. However, no statistically significant association was found between other obesity markers, including commonly used BMI, with MGUS risk.

Our results imply that the inconsistent findings from previous studies may be caused by using different obesity measures and selection of study participants.^3,15^ Although BMI is very popular in epidemiological studies for the diagnosis of obesity, the present evidence suggests that the impact of the BMI is variable and limited.^32^ The obese phenotype is multifaceted and can be characterized by measures of body fat and skeletal muscle mass.^33^ However, BMI is not considered a good proxy for fat mass. As noted, the Pearson correlation coefficient between BMI and %BF is only 0.58 in NHANES participants. Compared to BMI, BIA and digital anthropometry both have the potential to provide accurate measures of fat mass and distribution in clinical settings. Furthermore, this study suggests that monitoring body fat percentage using advanced digital anthropometry methods, such as DXA, provides more accurate and informative assessments than BIA for identifying individuals at higher risk of MGUS.

The mechanism by which increased fat percentage elevates the risk of MGUS may be multifactorial, involving various metabolic and inflammatory pathways.^34^ Adipose tissue, particularly visceral fat, is not merely a passive storage depot for excess energy but an active endocrine organ that secretes numerous bioactive molecules known as adipokines. These adipokines, including leptin, adiponectin, and pro-inflammatory cytokines, can create a chronic low-grade inflammatory state.^35,36^ And chronic inflammation is a well-known risk factor for the development of various malignancies, including hematologic cancers.^36-38^ Additionally, increased adiposity is associated with higher levels of insulin-like growth factor (IGF-1), creating a pro-oncogenic environment that may contribute to the initiation and progression of plasma cell disorders.^39^ Animal studies also have shown that diet-induced obesity is associated with the development of MGUS in wild-type mice, linked to increased levels of IGF.^40^

Our study suggests that lifestyle interventions targeting a reduction in body fat percentage, rather than focusing solely on weight loss, may play a critical role in preventing MGUS. Nutritional interventions focusing on balanced diets rich in fruits, vegetables, and whole grains, while reducing the intake of sugar-sweetened drinks, fast and processed foods, and red and processed meat, may help lower the risk.^41^ Rather than focusing solely on creating a negative energy balance through diet control and calorie reduction, strength training and resistance exercises, in particular, are helpful to increasing muscle mass, which not only helps lower fat percentage but also supports long-term metabolic health. Increasing physical activity levels has shown an inverse association with MGUS, suggesting that regular exercise can be a protective factor.^16^

The strength of this study includes the use of NHANES III and continuous NHANES, which are nationally representative data. We analyzed 7 obesity markers (6 out 7 are subjectively measured), instead using BMI only. The results of this study provide comprehensive analysis of the association between MGUS and obesity.

This study also has limitations. Firstly, there is a lack of progression information for those diagnosed with MGUS. Secondly, as with all observational studies, there is the potential for confounding data, which may affect the validity of our findings. Furthermore, the cross-sectional nature of NHANES data limits the ability to infer causality. More studies are needed to confirm these associations and elucidate potential mechanisms.

## Data Availability

All data produced in the present work are contained in the manuscript

## Notes

### Competing Interest Statement

The authors have declared no competing interest.

### Funding Statement

This study was funded by the National Cancer Institute of the National Institutes of Health under Award Numbers U01CA265735.

### Author Declarations

The study used ONLY openly available human data that were originally located at:https://www.cdc.gov/nchs/nhanes/index.htm

