## Supplementary Figure S1; Supplementary Table S1 for "Body Fatness Increases Risk of Monoclonal Gammopathy of Undetermined Significance in the U.S. Population"

**Appendix**

**Table S1:** The association between body composition and risk of developing MGUS using 5 imputed data from NHANES 1999-2004.

|  | **Multivariable-adjusted Odds Ratio [95% CI]** | | | | |
| --- | --- | --- | --- | --- | --- |
|  | Imputation 1 | Imputation 2 | Imputation 3 | Imputation 4 | Imputation 5 |
| ***All MGUS*** |  |  |  |  |  |
| Fat-free mass (kg) | 1.00 [0.97, 1.02] | 0.99 [0.97, 1.02] | 1.00 [0.97, 1.02] | 0.99 [0.97, 1.02] | 0.99 [0.97, 1.02] |
| Total body fat (kg) | 1.02 [1.00, 1.04] | 1.02 [0.99, 1.04] | 1.01 [0.99, 1.04] | 1.01 [0.99, 1.04] | 1.02 [1.00, 1.04] |
| Body percentage fat (%) | **1.04 [1.01, 1.07]*** | **1.04 [1.01, 1.08]*** | **1.03 [1.00, 1.07]*** | **1.04 [1.01, 1.07]*** | **1.04 [1.01, 1.08]*** |
| ***IgG/IgA MGUS*** |  |  |  |  |  |
| Fat-free mass (kg) | 1.00 [0.98, 1.03] | 1.00 [0.98, 1.03] | 1.01 [0.98, 1.03] | 1.00 [0.98, 1.04] | 1.00 [0.97, 1.03] |
| Total body fat (kg) | **1.02 [1.00, 1.05]*** | 1.02 [1.00, 1.05] | 1.02 [1.00, 1.05] | 1.02 [1.00, 1.05] | **1.02 [1.00, 1.05]*** |
| Body percentage fat (%) | **1.06 [1.02, 1.10]*** | **1.06 [1.02, 1.10]*** | **1.05 [1.01, 1.10]*** | **1.06 [1.02, 1.10]*** | **1.06 [1.02, 1.10]*** |

**p < .05. All logistic models were adjusted for demographic characteristics (i.e., age, gender, race, education level, and ratio of family income to poverty)*

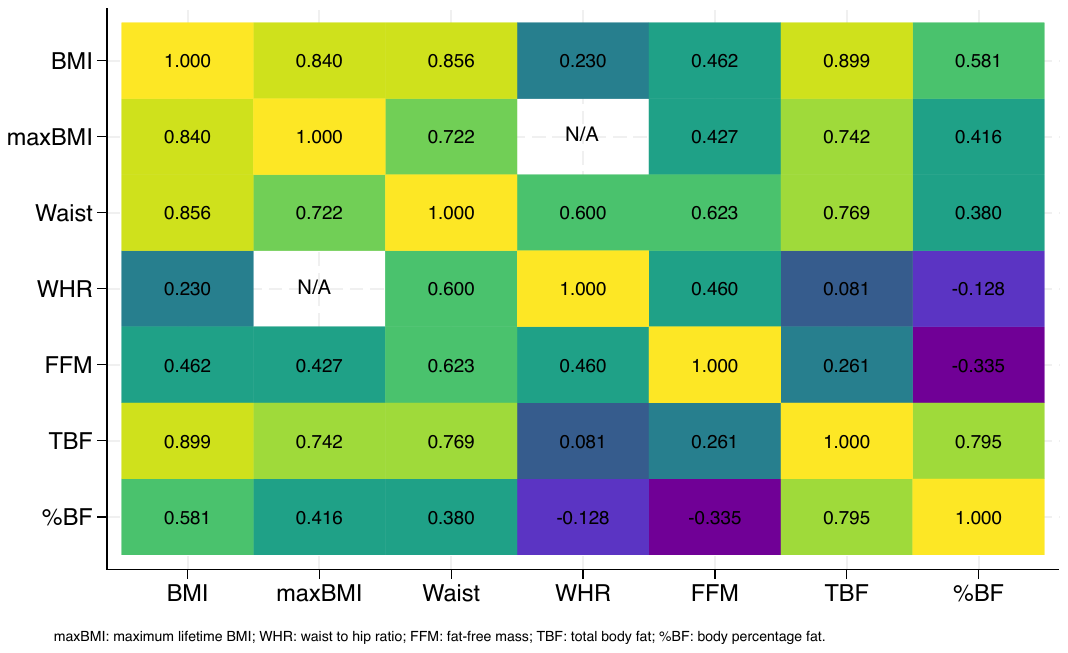

**Figure S1:** Correlations between obesity markers in NHANES III and NHANES 1999-2004.
